# Investigating the effects of healthcare access and level of education as risk factors for hypertension among male patients aged 35-70 attending government health centers in Kenya: A scoping Review

**DOI:** 10.1101/2025.05.04.25326761

**Authors:** Collins Kipkemoi Chepkwony, David Mulenga

## Abstract

Hypertension is one of the non-communicable diseases that is on the rise, also referred to as a ‘silent’ chronic killer. It is defined as a systemic blood pressure of more than 140mmHg systolic and more than 90mmHg diastolic. Education level and accessibility to health care are vital risk factors that have a direct impact on hypertension. Awareness enables an individual to diminish the myths and misconceptions about hypertension, which include the use of herbal medications for hypertension.

This review sought to evaluate the effects of healthcare access and education as risk factors for hypertension among male patients aged 35-70 who attend government health centers in Kenya.

Primary studies were reviewed using three databases (PubMed, Google Scholar, Scopus, and Science Direct). This was done systematically by employing the illustration by Arksey, O’Malley, and Levac. The scoping review adopted a broad literature search that allowed the transparency, reproducibility, and state of reliable literature. A narrative synthesis was used to describe the included studies and results.

Eleven primary studies were found to be eligible for this review. These articles met all the inclusion criteria. The studies were obtained from Western (n=3), Nairobi (n=2), Rift Valley (n=2), Coastal (n=1), Central (n=1), Eastern (n=0), North Eastern (n=0), and All regions (n=2).

The studies’ results picked out pertinent factors. Group medical visits are lower in both males and females, which influences disability-adjusted life years (DALYs). Screening also came out as another very signif cant reason for hypertension; impaired screening results in delayed diagnosis of hypertension.

Education has a direct impact on hypertension, i.e., the more educated one is, the lower the chances of becoming hypertensive. For male participants who took the screening positively and were started on treatment, the blood pressure was well achieved, and linkage to the nearby health facilities was done. Awareness about hypertension leads to early detection and as a result early initiation of therapy. Linkage to the facilities ensures that the clients are followed up keenly to avoid late treatment, encouraging the population to obtain health insurance is essential to making sure that they obtain access to health care.

## 1. Introduction

### Overview

This scoping review covers the effects of healthcare access and education as risk factors for hypertension. It discusses in detail the eleven studies that were eligible for this review. A narrative synthesis was used to describe the included studies and results. Education has a direct impact on hypertension. Awareness about hypertension leads to early detection and, as a result, early initiation of therapy. Linkage to the facilities ensures that the clients are followed up keenly to avoid late treatment.

Hypertension is defined as elevated blood pressure of diastolic ≥90 mmHg or systolic blood pressure of ≥ 140 mmHg above the normal WHO (2023). This non-communicable disease is the major contributor to morbidities and mortalities because of cardiovascular complications. (Bludorn and Railey, 2024).

Hypertension (HTN) hardly shows any symptoms except severe headaches in extreme situations; other than that, this makes HTN go unnoticed for several years, hence becoming a silent killer. The complications begin to manifest at a later stage, this is different when compared to other NCDs like diabetes, which have symptoms. Upon early diagnosis of hypertension, therapy was instituted. This will improve the quality-adjusted life (QALY) of the patient, and as a result, the productivity of the patient will be well appreciated.

Global statistics show that in the US, 46.7% of the adult population, equivalent to 122 million adults, of which 62.8 million were men and 59.6 million were hypertensive women, this still explains the reason for more men being hypertensive than women even in this review.

According to Global Disease Burden, in 2019, the number of deaths due to Hypertensive Heart Disease was 1.1567 million (95% UI 0.8598 million to 1.2786 million) among the measured population across all age groups. This accounted for 2.046% of the burden of all 369 diseases, an increase of 76.63% (95% UI 62.06–74.49%) Lu et al., 2024

The complications of hypertension include cardiovascular diseases (aforementioned), renal complications, ocular complications (retinopathy)

A study by Burnier et al. (2021) regarding hypertension healthcare beliefs entailed 3196 healthcare professionals; it became clear that antihypertensive therapy was directly related to the time the clinician took to give health education to the client. This brings to the spot the level of education as a risk factor for hypertension.

Kenya is not the only country battling with the burden of hypertension. Still, in the global picture, HTN is responsible for over 10 million deaths annually, which is equal to all deaths of infectious causes combined. A large number of these cases are found in Low-middle-income Countries (LMIC) where most cannot access hypertension treatment due to reasons like healthcare accessibility or even negligence because of not being educated to know the importance of adherence Piot et al., 2016).

In Kenya, Hypertension is among the top four leading diseases accounting for more than 50% of prolonged hospital admissions. This has significant economic losses because of cardiovascular diseases. A study in Kenya done in 2019 showed that 34.7% and 31.4% of women and men, respectively, were hypertensive. High disease burden causes a strain on the health system and comes at significant economic vs societal costs. Studies further reveal a strong relationship between socioeconomic status because of individual health behavior and hypertension (Gaiser et al., 2024).

The significance of this review is drawn from the fact that healthcare access and level of education are pertinent risk factors for hypertension. According to the 2018 UN-Habitat report on Kenya’s profile, 27% of the population lives in urban areas, which means the larger proportion of 73% lives in rural parts. The urban growth rate is 4.23%. Rural areas have poor access to healthcare. Still, most of the people living in urban areas are educated, and hence, they have better health-seeking behavior as compared to those in rural areas. In some cultural aspects, some cultures still believe that hypertension is the disease of the rich, while the younger age group (35-40 yrs.) believe that hypertension is not their disease.

Furthermore, there is an increasing trend in Kenya among males who smoke, take alcohol, and smoke cannabis among other substances. Men are on the rise more into these substances than females. This speaks volumes on why more men are hypertensive, especially looking at the risk factors for hypertension. A study by Joshi et al, 2014 showed that of the 2045 subjects whose blood pressure was taken, 61.8% were hypertensive males while 56.6% were females.

In regards to healthcare access, the Kenyan Government has introduced the Social Health Insurance Fund (SHIF, which replaces the old funding Model, the National Health Insurance Fund (NHIF). Looking at SHIF has a wider array of benefits for clients with chronic illnesses and non-communicable diseases. In this case, for instance, hypertensive patients will be able to access medications from all public facilities and even seek medical attention from level 4-6 hospitals. This effort by the Kenyan government aims at ensuring that healthcare access to patients is achieved. As of September 2024, this funding model took effect despite the small challenges here and there; this health insurance model will be a change maker to the chronic diseases prospect.

It makes it vital to undertake this scoping review to address the global concern of this non-communicable disease; Hypertension. It is a clarion call to all the stakeholders to address hypertension preventive strategies to reduce the effects of its dire complications.

## 2. Methodology

### 2.1 Identifying the research question

The question of this scoping review is “What are the effects of healthcare access and education as a risk factor for hypertension among male patients aged 35-70 years attending government health centers in Kenya?

### 2.2 Identifying relevant studies

A search using three electronic databases was used; these are Google Scholar, PubMed, Scopus, and Science Direct.

A systematic literature review on the effects of healthcare access and level of education as a risk factor for hypertension among male patients visiting government health facilities in Kenya was performed. The studies bring out the use of scoping review as well illustrated by Arksey and O’Malley, 2005 and Levac et al.

This Scoping review adopts a broad strategic search as it allows the transparency, reproducibility, and state of reliable literature. Keywords have been used while searching for the literature, original peer-reviewed articles published in English The date limit was intentionally left open to ensure that there is a collection of all the relevant articles hence ensuring that there is a richness of data to answer the research question. The articles were obtained via an electronic database search such as Google Scholar. Pub med, Medline, and Scopus. Unpublished work was not included to ensure that there is quality of the findings. Medical peer-reviewed journals, public health, and social sciences journals were carried out extensively on the search database mentioned above.

The initial search utilized a broad search strategy whereby free text terminologies, synonyms, and headings related to the effects of education, healthcare access in hypertension, and males between 35-70 years. Then on the final bit, a general search combining the terms “effects of healthcare access in hypertension”, “ education risk factor for hypertension“:” high blood pressure in Kenyan males” “ “health centers attendance males in Kenya” and “effects of elevated blood pressure”.

The reference list of the obtained primary articles were further checked just for any availability of other potential studies that may be relevant to this scoping review. This is very essential as it allows a wider scope for the literature search and since there is a wider scope of the open data search as well as enabling more data to be obtained.

### 2.3 Selection of relevant & reliable studies

By ensuring that the eligibility criteria (inclusion & Exclusion) are taken into account. The study selection process was engaging as it entailed a literature search, search strategy refining, and articles review to ensure that they met the inclusion criteria.

For the studies to be included had to focus on male patients aged 35-70 years, the research papers, articles, and journals that address healthcare access and education level as risk factors related to hypertension in Kenya. Ensuring that both qualitative and quantitative studies are taken into account.

While the studies that were nonspecific to Kenya or even lacked data on hypertension risk factors were excluded. Additional studies published in a language other than English were excluded. In a scenario where let us say multiple articles reported the same data, only one paper was included to avoid duplication. The study yielded 804 searches.

### 2.4 Charting the data

This is the initial step in summarizing and synthesizing the evidence obtained from the search. This process was quite interactive since it entailed discussion and reflection across the authors. Keeping in mind that the principles set in place by Gough, Thomas, and Oliver were strictly adhered to. Gough et al., (2017) and Thomas et al., (2017).

The data charting form was developed using an Excel sheet. The data extracted included the title of the study, objectives of the study, authors’ publication year, the country where the study was conducted, study design, study population phenomenon key findings that relate to the review question.

### 2.5 Data synthesis and analysis

Thematic analysis was done on the extracted data; themes identified, and appropriate categories that clearly describe the breadth of the review question. Findings obtained from the thematic analysis were put in a table format and synthesized through a narration.

### 2.6 Collating, summarizing, and reporting the results

Following the recommendation by Levac et al., (2010), a narrative synthesis was used to discuss the included studies. Healthcare access and level of education were compared and the definitions were obtained, taking into consideration how various authors gave the description. This was done purposely to check for any commonalities or differences. Based on the two risk factors for hypertension (Healthcare access and Level of education), they were further classified based on the male PA patients aged 35-70 years and patients attending government health centers. Through this interactive process, evidence from the articles was extracted. This further allowed them to ensure that the wide array of narrative synthesis is well achieved.

### 2.7 Ethical approval

There was no direct involvement of any primary patient in this review, so institutional review board approval was not required.

### 2.8 Search outcome

Figure 1 below clearly explains how the search process was attained via the databases and other sources. 804 titles were identified as per the following databases:

1. PubMed
2. Scopus
3. Google Scholar
4. Science Direct

**Fig. 1.**
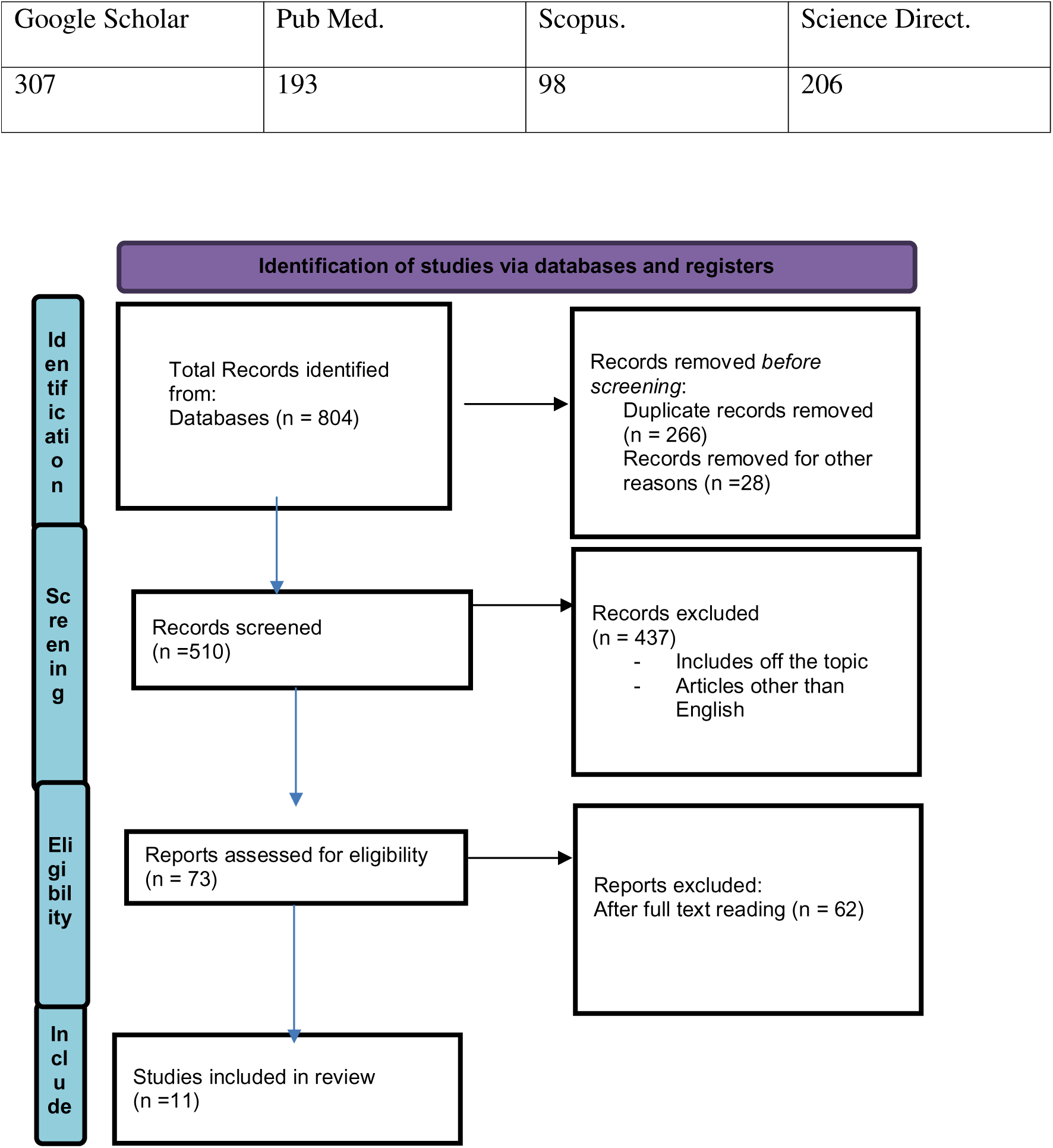
PRISMA 2020 flow diagram for the study selection process.

Articles that remained after the duplicates were removed were 510. Further assessment in terms of the screening was done, making the articles that remained after the screening to be 73 eligibilities obtained, and based on inclusion and exclusion criteria, 11 studies were included in the scoping review.

## 3. Results

### 3.1 Identifying potential studies

The electronic databases that were used in this review were Google Scholar, Scopus, PubMed, and Science Direct. Each yielded the following results:

Google Scholar – 307, PubMed – 193, Scopus – 98, Science Direct - 206

These databases yielded 804 searches. The process of searching for articles was very intense, entailing four key steps. Firstly, the identification stage, where there is a wider online search on the above-mentioned databases, ensures that the appropriate keywords are entered appropriately to give a broader search yielding 804 searches. The second stage is where the article is filtered depending on the title and abstract and where the duplicates are filtered, giving 510 searches. Further, the eligibility criteria (Inclusion and exclusion) were thoroughly filtered, narrowing the searches to 73. Then the last step is where the studies included are captured and a detailed report on each of the studies is discussed, the included studies were 11 in my final data extraction.

### 3.2 Characteristics of the studies included

The peer-reviewed literature on healthcare access and the level of education as risk factors of hypertension. The majority of the articles were obtained from all the regions in Kenya, which include Nairobi, Western, Rift Valley, Nyanza, Central, coastal Eastern, and North Eastern parts of Kenya. Among these articles, it addresses the level of education. The studies reported were as follows.

The table below presents a full range of the included studies; the different studies examined how healthcare access and level of education are greater risk factors for hypertension in men. Various studies further go ahead to ensure that a vivid description is given of why men are at greater risk as compared to women. Together, the articles reported on how healthcare access and level of education are the greatest risk factors for hypertension; this strongly gives a reason why HTN is one of the NCDs that is on the rise in Kenya. The numbers of the studies are Nairobi (n = 2), Western (n = 3), Rift Valley (n = 2), Nyanza (n = 0), Central (n = 1), coastal (n = 1) Eastern (n = 0) and North Eastern (n = 0). All regions (n=2). There was no specific range for the search period to allow rich data search; the articles obtained were of recent publication. Most studies applied quantitative data collection methods, while mixed methods were also used in some selected articles.

### 3.3 Level of education and healthcare access as a risk factor for hypertension

Various factors have been elicited in the articles regarding how each of the aforementioned factors hurts HTN. According to Joshi et al. (2014). The slums have a higher number of populations ailing from hypertension and its resultant complications due to poor health care access. This results in higher cases of mortality. The results from the articles are quite distinctive, painting a picture that an extensive public health strategy has to be made to ensure that these risk factors are greatly addressed.

### 3.4 Results presentation in tabular form

The results of the 11 included sources are listed down capturing the name of the author, year of publication, the title of the article, the publisher, and the results of the articles.

**Table 1.**
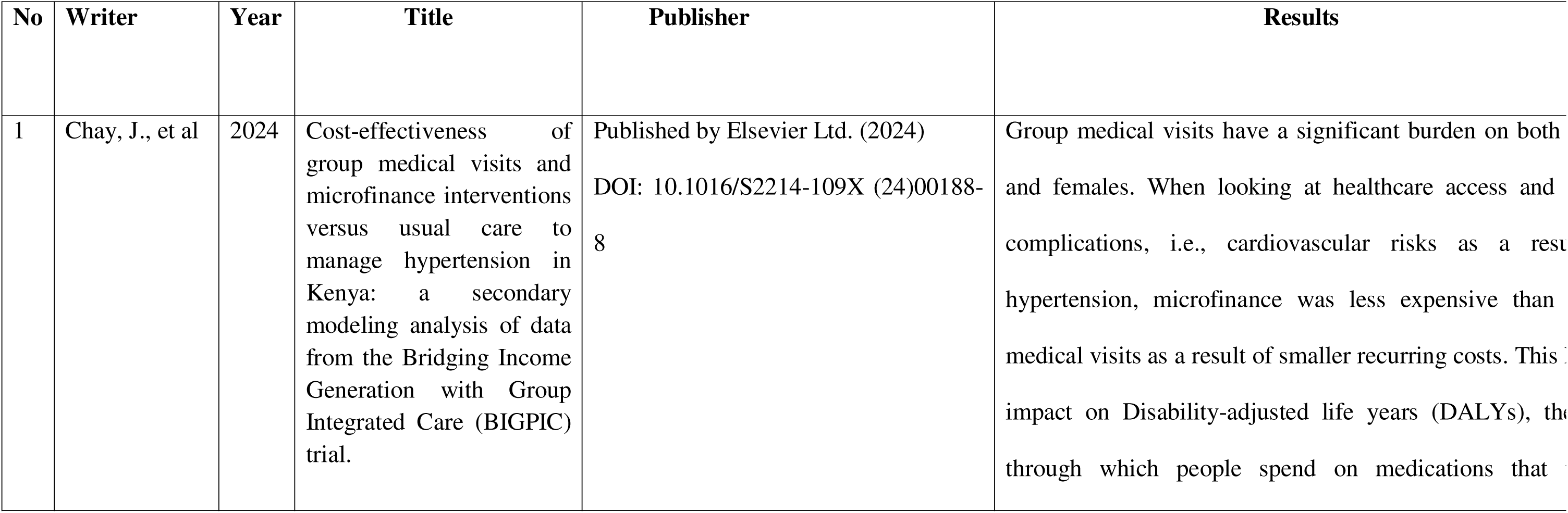
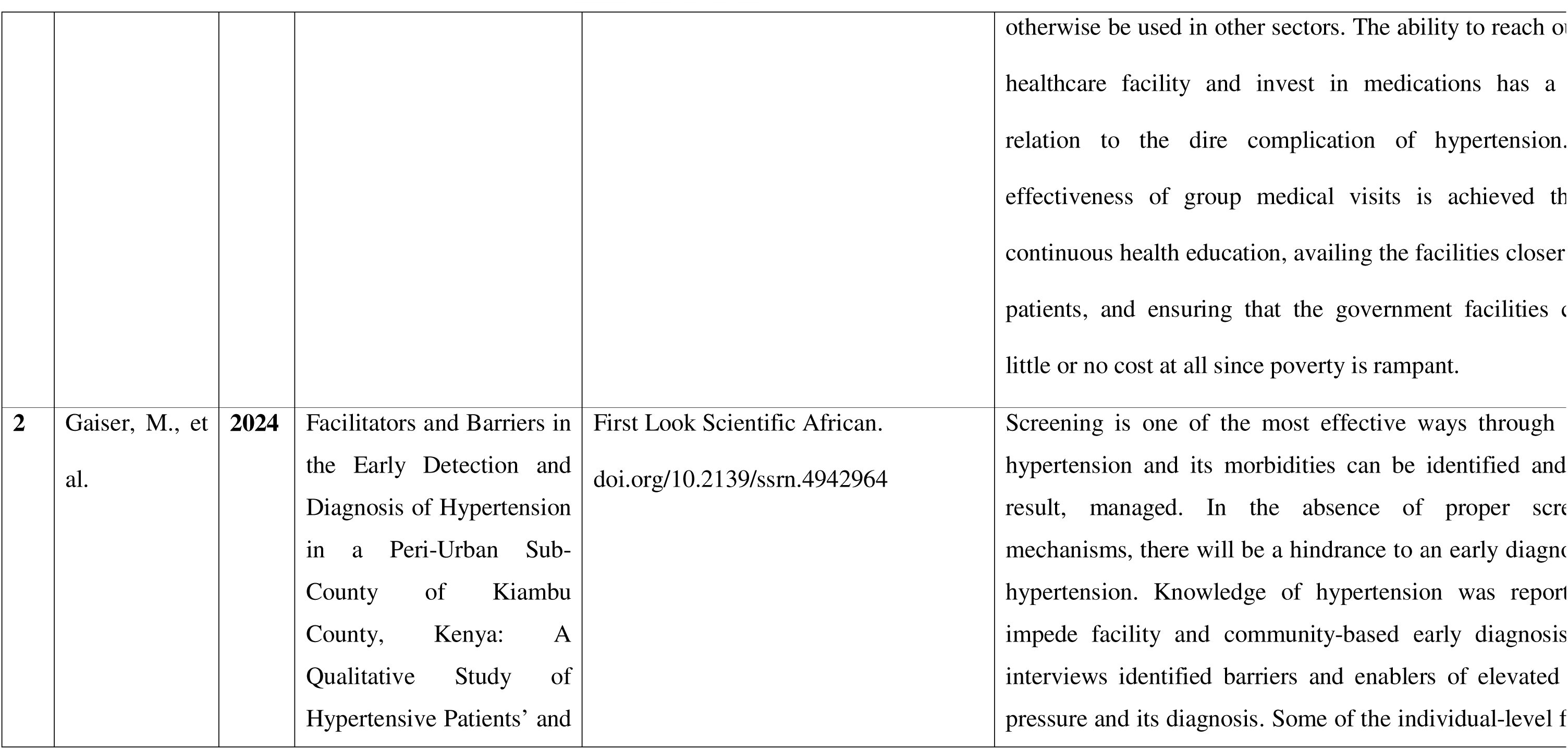
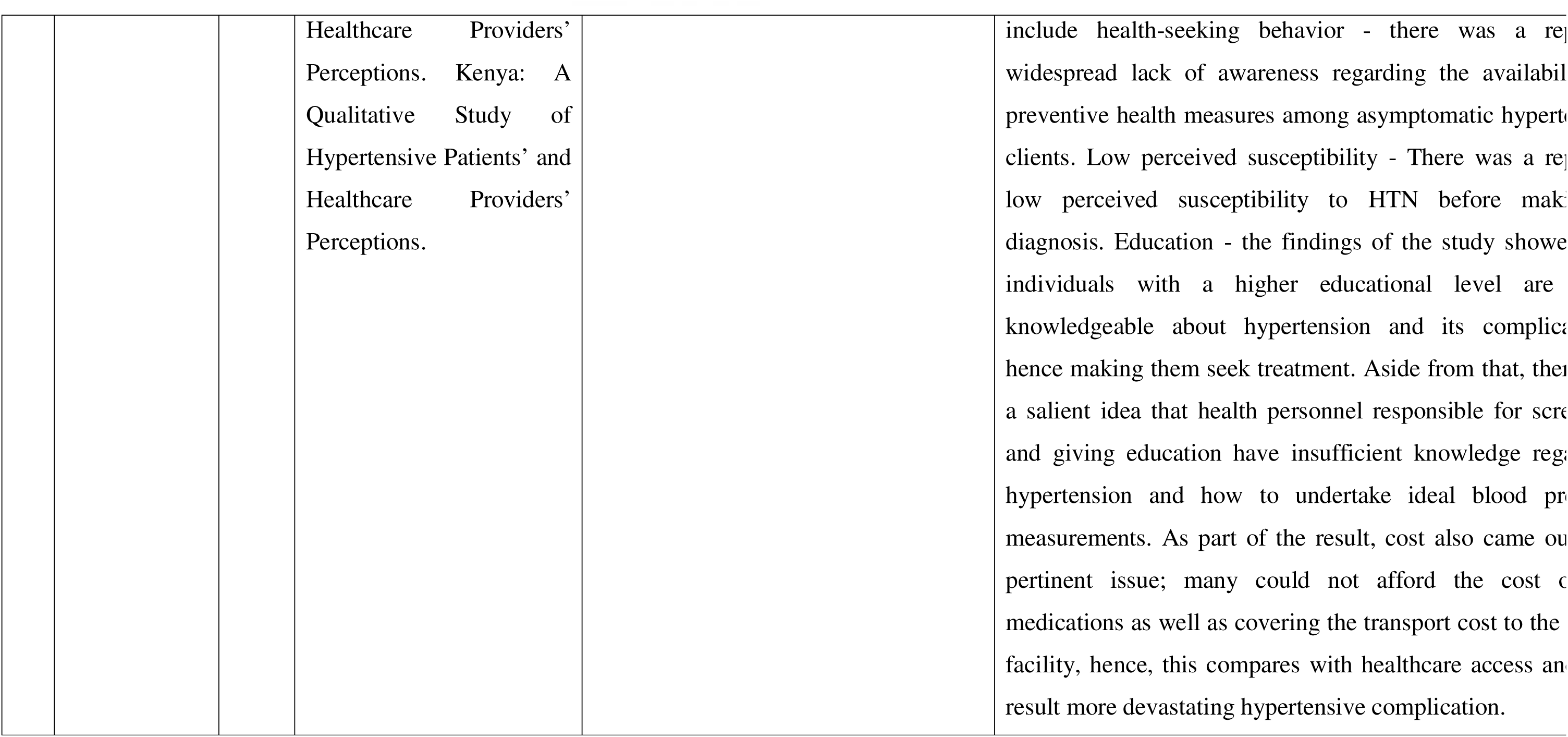
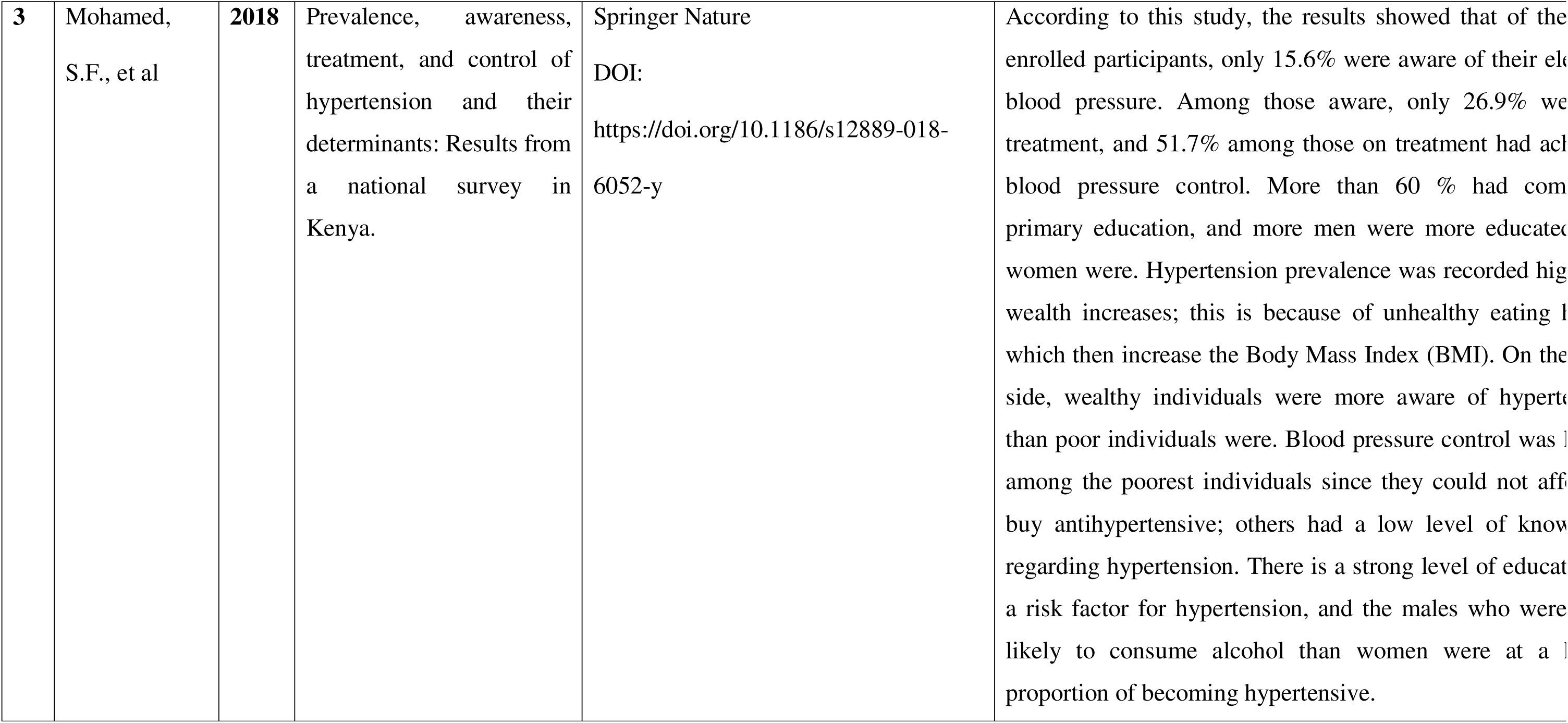
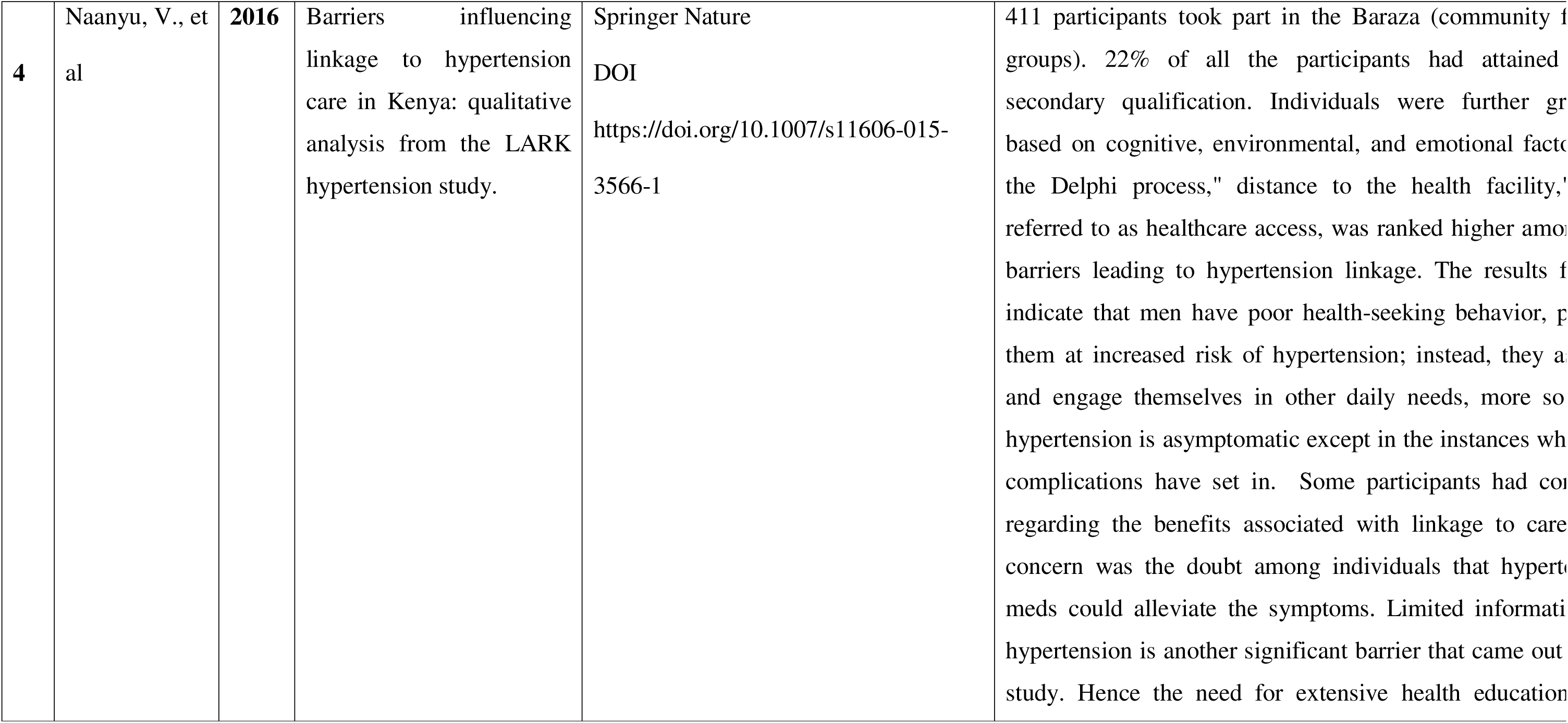
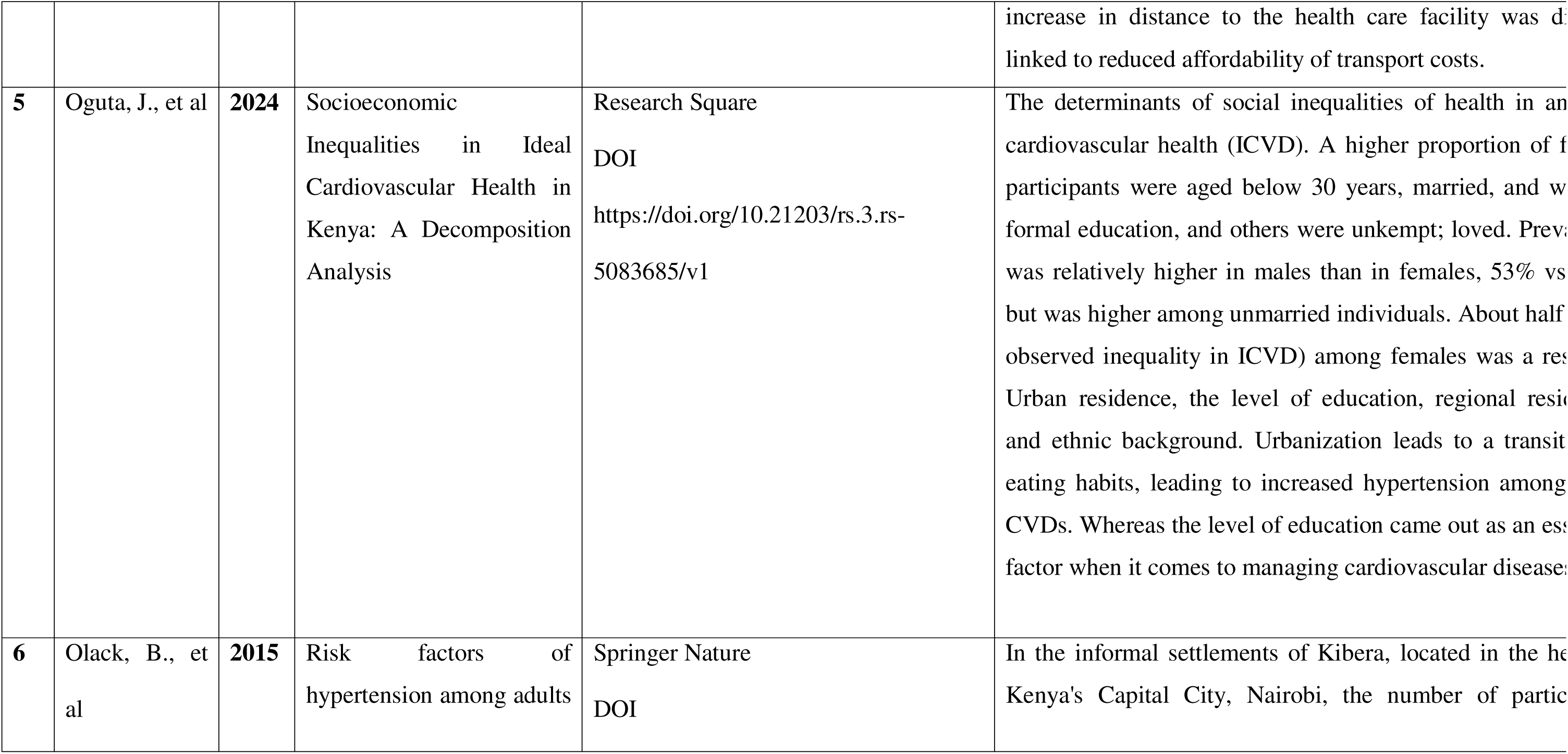
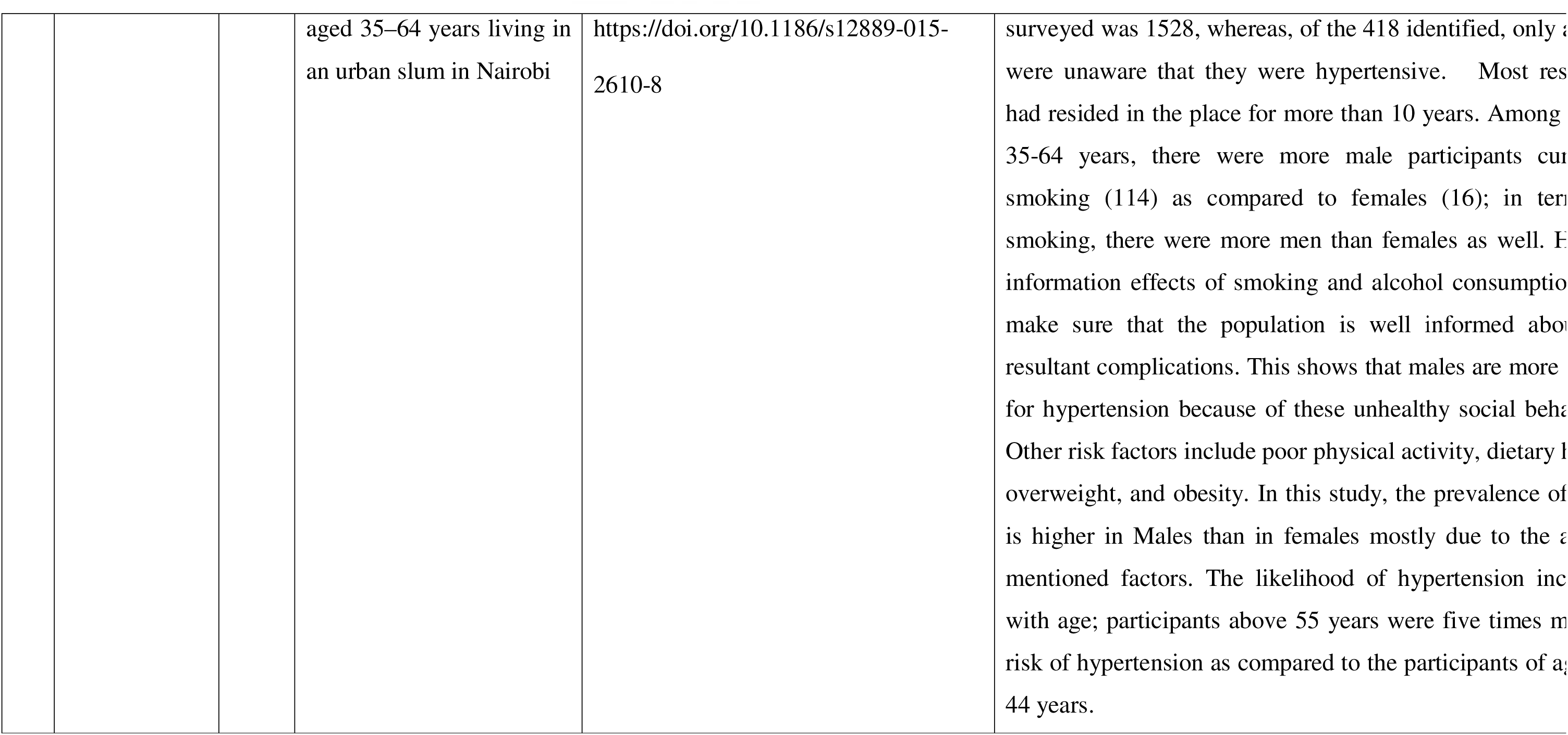
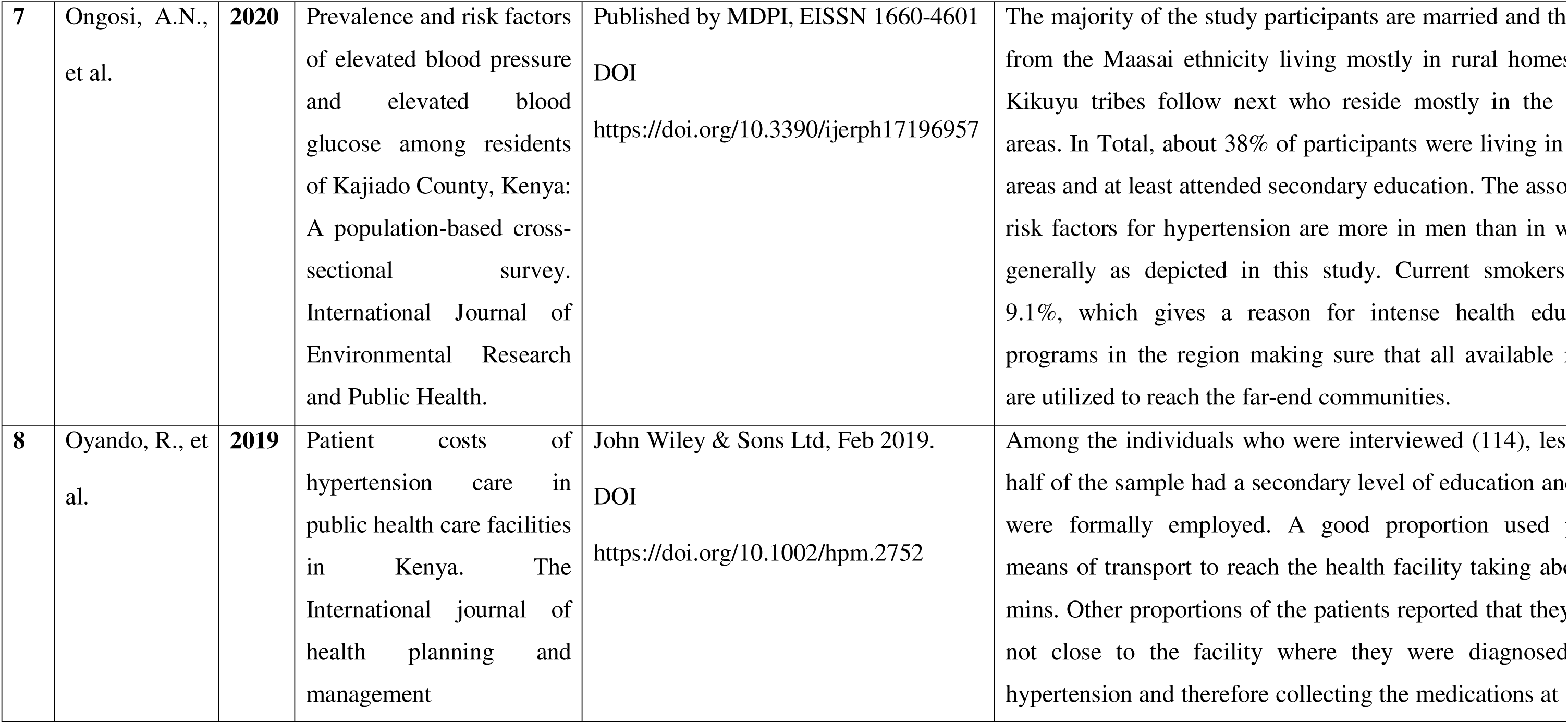
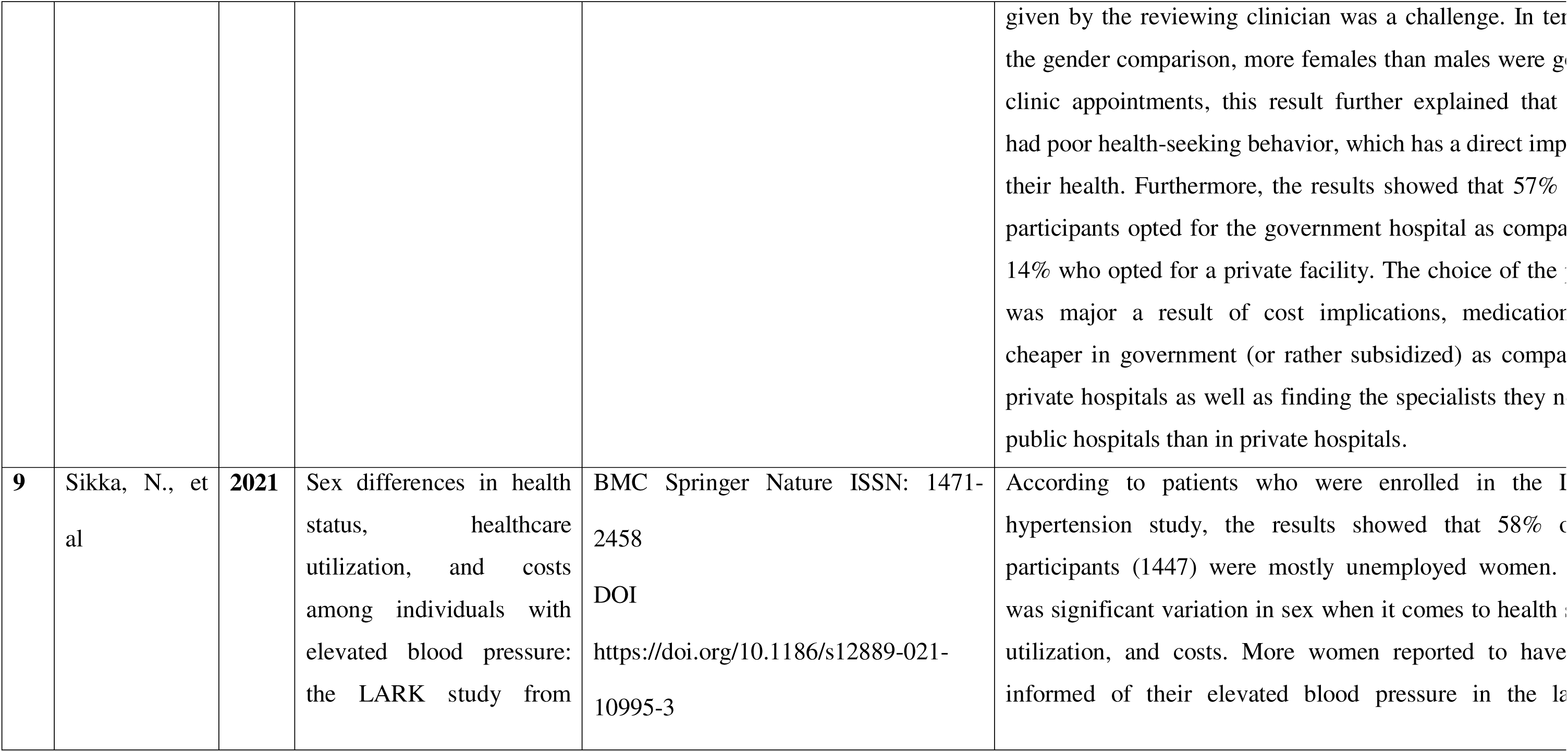
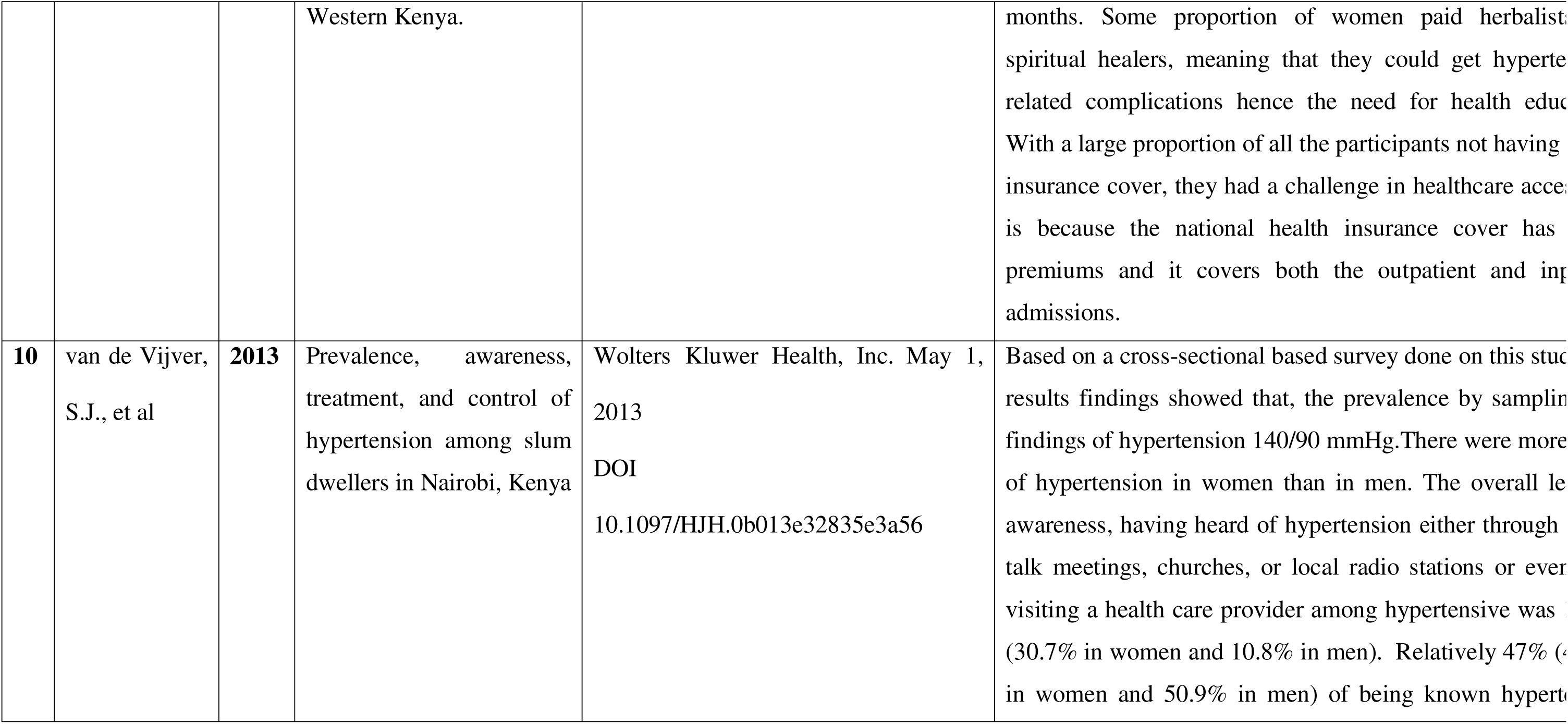
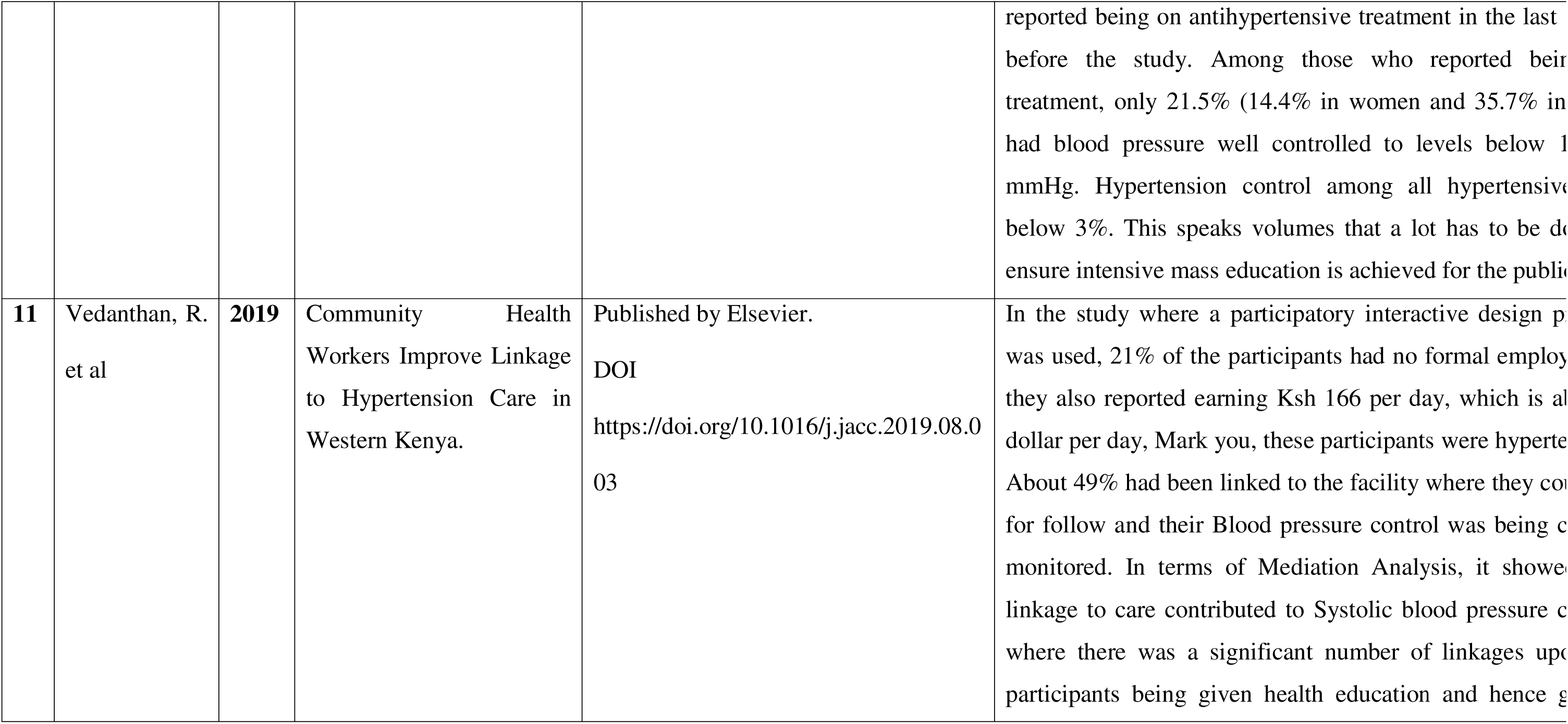
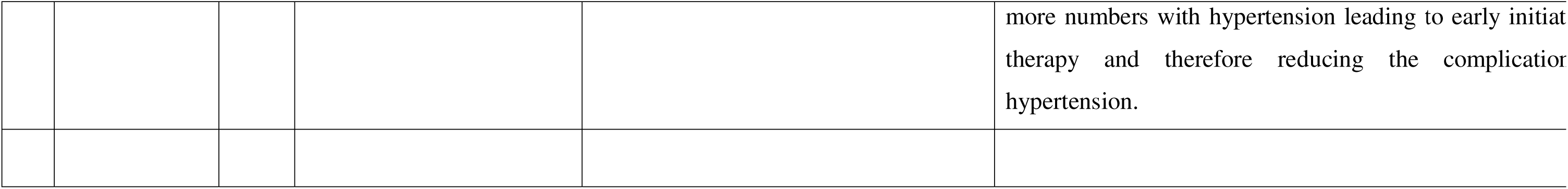
shows the results.

### 3.5 Results Synthesis

The research result by Chay, et al (2024) illustrated that continuous medical education is very vital to the population to reduce the cost through which people spend on buying drugs; instead, the finances are channeled to another economic advancement.

Gaiser, et al (2024) also went ahead to mention that early diagnosis is very vital for hypertension this will ensure that the complications that would have occurred are prevented. Despite the lack of awareness regarding the availability of preventive health measures among asymptomatic hypertensive clients. The study also finds out that the cost of both medications and transport was a great concern since this impairs healthcare care access resulting in more hypertensive complications like CVD.

The study by Mohamed, et al (2018) showed that Hypertension prevalence was recorded highly as wealth increases, this is because of unhealthy eating habits, which then increase the Body Mass Index (BMI). On the flipside, wealthy individuals were more aware of hypertension than poor individuals were. Blood pressure control was higher among the poorest individuals since they could not afford to buy antihypertensive, others had a low level of knowledge regarding hypertension.

The search result by Naanyu, et al (2016) clearly described the Delphi process; the distance to the health facility was ranked as one of the leading barriers to hypertension linkage. The results further indicate that men have poorer health-seeking behavior than women do. Limited information about hypertension among the participants was a concern.

The study by Oguta, et al (2024) found that prevalence was higher in males than females among the participants who had no formal education and were unemployed as well. Level of education, associated with regional residence, and eating habits had a significant role in being at risk of Hypertension and other non-communicable diseases.

Moreover, Olack, et al (2015) findings were very classical that in the informal settlement, Kibera located in the capital city of Kenya, among the participants who were surveyed, more males than females were smokers and alcoholics, and most of them had resided in the place for more than 10 yrs. The interesting bit is that most of them had no awareness about the effects of smoking and the use of illicit alcohol brew; they had no connection between the health effects of smoking, or alcohol intake to smoking. This paused a challenge that there is a need for intense health education for the locals.

Ongosi, et al (2020) this study focus on the Maasai ethnic group who reside in the eastern part of the country, Maasai are known for their reserved culture. There is an extensive use of nonscientific means of medications for their ailments; most of the participants were smokers hence the need for awareness to improve on their lifestyle changes since this will hurt their health.

According to Onyango et al (2019), the results showed that less than half of the sampled population had secondary education. Most of the participants opted to go to the government health facility since it was closer to them and the cost of buying the medications was cheaper than in private facilities.

Aside from that, the results of the study done by Sikka et al. (2021) showed that there was significant variation in sex when it came to health status, utilization, and costs. More women reported having been informed of their elevated blood pressure in the last 12 months. A large proportion of the participants did not have health insurance coverage, hence impairing them from having healthcare access.

Moreover, the study by van de Vijver et al. (2013) summed up the fact that hypertensive control is still very low, and there is the perception that hypertension is the “disease for the rich” Intensive mass education is paramount so to address the myths and encourage linkage to the facility.

Finally, Vedanthan, et al (2019) on the study found that linkage to the facility for close follow-up is key. This enabled early diagnosis of hypertension hence initiation of the treatment leading to reduced complications associated with hypertension.

## 4. Discussion

This scoping review is very essential in terms of critically analyzing the effects of the level of education and healthcare access as a risk factor for Hypertension, the analysis of the 11 articles gave an in-depth understanding of hypertension in regards to the Level of education and Healthcare access as the most profound risk factors. The discussion is classified into individual level, community level, Policy Levels, and strengths and limitations. The findings are valuable in terms of eliciting factors surrounding the risk factors for hypertension. It also came out clearly in the studies analyzed that spiritual factors, cultural factors, and the use of herbal medications were some of the factors leading to hypertensive complications.

### 4.1) Individual factors

The study found that there was limited awareness and several other myths and misconceptions regarding hypertension diagnosis. Consistent with the findings in this review, a lack of knowledge among participants was noted in six of the reviews making it one of the greatest risk factors. This is not only consistent in Kenya but also among other Low and middle-income countries (LMIC). The studies also reported that hypertension was reported with increasing age, sometimes referred to as a lifestyle disease. Other individual factors reported include misconceptions and the use of herbal medications. Being knowledgeable enables one to give credit to scientific medications since it has been researched and tested but convincing the illiterate to accept that is an uphill task.

It was further described clearly in the studies that men have poor health-seeking behavior; they only go to the hospital in the advanced stage of the disease, hence hurting their health. Men also are more engaged in behaviors like smoking and alcohol consumption, which puts them more at risk of hypertension and its resultant complications.

Other social demographic aspects like age, gender, level of education, and socioeconomic status were linked to having a closer influence on hypertension on gender parity; women were strongly recognized to have a greater uptake of screening services, though the reasons were not adequately discussed in this review.

Still, in the studies, it also came out clearly that acceptance is one important aspect. Most participants below 45 years old believed that hypertension is only the disease of the old; they thought that their age was not an important consideration, and they feared being on lifelong medications and, more so, other cardiovascular complications.

### 4.2) Community Level

According to the studies in this review, family, and friends influence seeking medical attention, which is considered a promotive approach. Healthcare access was one of the factors that influenced treatment, the study by Sikka, et al (2021) pointed out that healthcare access could be a big hindrance to combatting hypertension. In the Eastern and North Eastern parts of Kenya which are arid areas the accessibility of the healthcare facility is a concern, previously the presence of militia groups destabilized the security further affecting accessibility to health facilities in the region. Studies also mentioned that places of worship like churches were a good avenue to give education on hypertension screening diagnosis & risk factors since most of the community members have an affiliation to at least one place of worship. Religious leaders have a significant social status and influence in the community.

In addition to that, the study by Sikka et al (2021) brought out the concern about herbal medication use; some participants opted for herbal medications and traditional healers to be “healed” of hypertension. This has a great impact on oneself and the community at large. Health education had to be intensified to ensure that the community could have a clear distinction between herbal medications and scientific medicines. Access to healthcare is ideally considered one’s right, This review picked out the fact that financial constraints impaired one’s ability to seek treatment for hypertension, and poverty was exacerbated by hard economic times.

### 4.3 Policy stakeholders

Having a strong and working public health system is paramount to ensuring that there is an active and functional community outreach, outreach activities, and designated hypertension facilities. The findings in these studies indicate that there is limited funding for public health, which impedes the functionality of many programs. This is further affected by the embezzlement of funds by various public organs. Rooted corruption derails the efforts of adequately addressing hypertension and its resultant complications like cardiovascular diseases, retinopathy, and renal failure.

Routine screening and health education on hypertension were considered a major facilitator of the detection of hypertension in this review. Collective responsibility and ensuring that proper linkage to the hypertensive clients is achieved.

**Fig 2.**
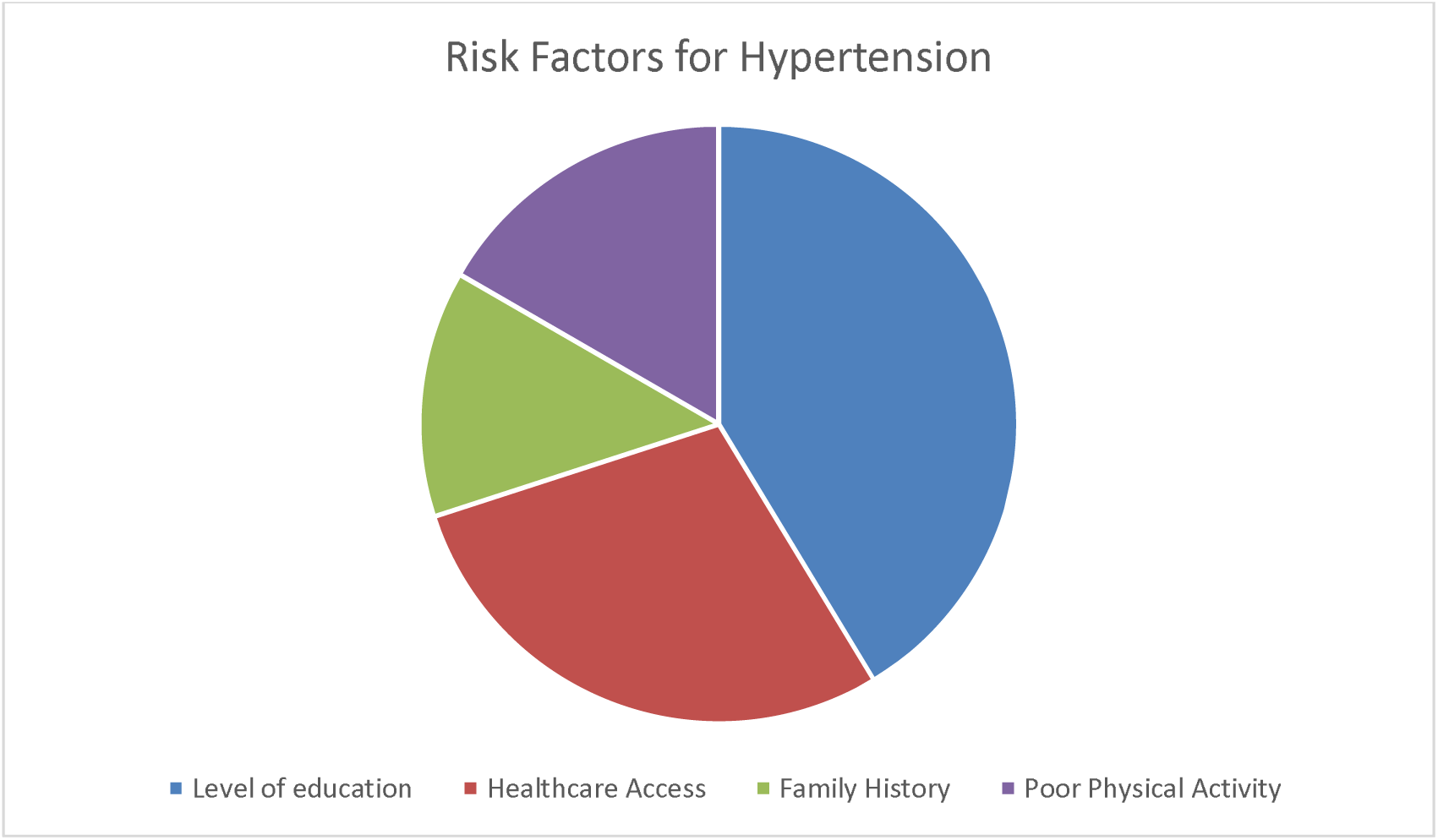
The pie chart below illustrates the various risk factors for hypertension.

### 4.4 Strengths and Limitations

The strength of this review is that it analyses the recent studies on the education level and healthcare access as risk factors for hypertension. The strength of this review includes the inclusion of awareness of hypertension among the participants, males being most at risk of hypertension, and how impaired healthcare access has an effect as a risk factor for hypertension. On the other side, the limitation that was elicited was that there are limited studies related to the review topic, but managed to obtain relevant studies. The exclusion criteria included any studies outside Kenya, Additionally; the treatment modalities for hypertension were not analyzed.

## 5. Conclusion

Hypertension is a non-communicable disease that has devastating effects. The complications are very dire and include heart failure, stroke, kidney complications, and retinopathy, among others. The level of education and healthcare access are the key risk factors for hypertension, they affect a particular individual. The more educated an individual is, the more they are responsible for their health. Hypertension is more common among the less educated as compared to the educated ones, and awareness is crucial for combatting the devastating complications of hypertension. Accessing health facilities could be hindered by distance to the facility, and poverty could impair one from paying for medications. Hypertension is a silent killer, and extensive health education through all the available media platforms is paramount. Information is power; that is why reaching out to as many people as possible is essential.

Utilizing formal and informal meetings in the community, Churches and other places of worship where numerous people gather should be an opportune place to convey health-related messages on Hypertension. This also gives the platform to demystify the myths and misconceptions as well as the use of herbal medications and visits to traditional herbalists/healers for treatment for hypertension. Males are more at risk than women because of poor health-seeking behavior in men compared to women. Men tend to visit the health facility when the complications become severe and they impair their normal functioning, i.e., gradual vision loss because of hypertensive retinopathy. Men engage themselves more in unhealthy behaviors such as smoking and alcohol consumption, which put their health at risk for hypertension. It is a collective responsibility for every one of us that lives in a healthy community free of preventable/ avoidable non-communicable diseases like hypertension. The government is spending substantial resources on this menace in the management of the complications, which could otherwise be avoided by altering our lifestyles.

## 6. Recommendations

### 6.1) Health education

Promoting awareness of hypertension is a minimum. All the necessary stakeholders from the community to the county and then to the national level should be engaged. This should make this a collective responsibility.

### 6.2) Hypertension screening

This will enable early detection and diagnosis, which will ensure timely initiation of therapy and hence close follow-up, reducing the latter complications of hypertension.

### 6.3) Further Research

More Studies and scoping reviews to be done around this topic on risk factors for hypertension, this will enable continuous realization of gaps that need to be investigated. Furthermore, international partners to support any studies related to this area since this is a global concern.

## Data Availability

I make fully available all the data that i obtained in my paper.

